# Early evidence of a higher incidence of COVID-19 in the air-polluted regions of eight severely affected countries

**DOI:** 10.1101/2020.04.30.20086496

**Authors:** Riccardo Pansini, Davide Fornacca

## Abstract

COVID-19 has spread in all continents in a span of just over three months, escalating into a pandemic that poses several humanitarian as well as scientific challenges. We here investigated the geographical character of the infection and correlate it with several annual satellite and ground indexes of air quality in: China, the United States, Italy, Iran, France, Spain, Germany, and the United Kingdom. Controlling for population size, we found more viral infections in those areas afflicted by high PM 2.5 and Nitrogen Dioxide values. Higher mortality was also correlated with relatively poor air quality. In Italy, the correspondence between the Po valley pollution and SARS-CoV-2 infections and induced mortality was the starkest, originating right in the most polluted European area. Air pollution appears to be for this disease a risk factor similar to smoking. This suggests the detrimental impact climate change will have on the trajectory of future respiratory epidemics.

## Introduction

From the first detected **outbreak** of a new member of the coronavirus (CoV) family^1^ in Wuhan, Hubei Province, China^2-4^, SARS-CoV-2^5^ has rapidly spread around the world^6^, with governments and institutions showing mixed results in its effective containment^7,8^. Certain regions have been much more adversely impacted in terms of the rate of infections and mortality rates than others, and the full reasons for this are not yet clear. This paper shows preliminary, yet compelling evidence of a correlation between air pollution and incidence of COVID-19 in eight countries.

Air pollution is notoriously known to cause health problems and, in particular, respiratory diseases, to individuals exposed for longer than several days per year^9-13^. Moreover, pollutants in the air which get absorbed systemically are significant underlying contributors to the emergence of respiratory viral infections^14^ including the previous SARS-CoV-1^15^. Air pollution has been shown to be strongly associated with a high incidence of other respiratory infections^9,12,14,16-23^ and higher mortality rates^10,11^. In particular, PM 10 and PM 2.5 have been linked to respiratory disease hospitalisations for pneumonia and chronic pulmonary diseases^16-22^. The ACE2 receptors of bronchial, alveolar, interstitial and other pulmonary cells can undergo to a state of chronic cellular inflammation due to air pollution and, concurrently, SARS-CoV-2^24^. There is some further experimental evidence that emissions from diesel and coal affect the lungs, causing pathological immune response and inflammations^25,26^, voiding past disputes^27^ that only high concentrations of these gasses are needed to cause pathologies.

A number of airborne microorganisms can directly infect other people’s mucosae or travel further into the air and onto surfaces causing delayed infections. The **particles** of several pollutants such as PMs and NO_2_ can act as a vector for the spread and extended survival in the air of bioaerosols^28-33^ including viruses^34-38^ such as measles, the avian flu H5N1, and the syncytial virus. A first hypothesis in this direction, not examined here, has arisen for SARS-CoV-2 in Northern Italy^39,40^.

The strong **containment** measures adopted firstly by the Chinese government have necessarily biased the natural virus spread^41,42^, not allowing the virus to distribute evenly across the country’s territory. Recent findings have shown that non-pharmaceutical interventions such as lockdowns significantly decreased the transmission of the virus in Europe^43^. What shall be noted, though, is that its appearance was recorded in a Chinese area affected by some of the highest air pollution in the world, and it showed a relatively high virulence there. In the case that, as in Italy^44^, the onset of the infection went undetected for weeks before the outbreaks became apparent, air pollution might have played a more relevant role in the exacerbation of the virus.

Some personal risk factors (also discussed in S.I.) have been associated with higher morbidity and mortality of COVID-19, including male gender and having hyperactivated airways receptors of both air pollution and smoking^45-47^. A high **population density** boosts the virus spread, but taken alone it should not be a reliable predictor for a higher virulence and a higher mortality^48^. Another evident predictor variable is **transportation**. The surrounding areas of transport hubs such as airports and large train stations should witness the appearance of the virus earlier than other less connected zones increasing its transmission^42,49-52^. (Further risk factors introduced in S.I.)

One factor that needs further investigation is the role of long-term exposure to **air pollution** in the spread of COVID-19 and possibly higher mortality rates. Long-term or chronic exposure, is a continuous or repeated contact with a toxic substance over a long period of time (months or years), and it is expressed by annual averaged data^53^. We therefore expand upon the very first study of this kind later followed by others, which we released in early-April, looking at this phenomenon in three countries^54^. By controlling for population size and density, here we investigate whether there is a correlation between long term exposure to air pollution and SARS-CoV-2 causing respiratory diseases in second order level administrations of eight countries: China, the United States, Italy, Iran, France, Spain, Germany, and the United Kingdom. Our **hypotheses** were: (1) is there a higher incidence of COVID-19 infections in areas chronically afflicted by poorer air quality?; and (2) is there a higher COVID-19 mortality rate in these highly polluted areas?

## Methods

We selected eight countries particularly affected by the virus and evaluated the potential correlation between air quality metrics and infections at the finest granularity available. Results for each country were analysed separately, controlling both COVID-19 and air pollution variables for potential relationships with population densities as well as the presence of the bivariate virus / pollution spatial clusters. (More on this in S.I.)

### Data collection and processing

The COVID-19 dataset (updated till the end of May) was compiled at the second-level administrative subdivision level (U.S.A. counties equivalent), updated until around the end of May; however, a few geographical and time adaptations were required for some contentious administrations which do not make public all the data (the details in S.I.).

Both infections and deaths due to COVID-19 were collected and normalised by population size per administration unit (100,000 residents). Population densities for each unit’s area were calculated at 1 square km resolution.

Air quality information was retrieved from long-term satellite observations and averaged at the administrative unit level. Additionally, ground measures for the United States, China and Italy were collected from various sources (see S.I.). Combining all these measures poses compilation challenges^55^. Satellite data hold several advantages over ground station data, such as regular and continuous data acquisition, quasi-global coverage, and spatially consistent measurement methodologies. On the other hand, ground stations offer real measures of single pollutants instead of deriving it from spectral information; however, they require more or less arbitrary estimations (such as interpolation) to fill spatial gaps. (More in S.I. discussion)

Table S1 in S.I. summarises the datasets used.

### Data Analysis

Exploratory analysis of the variables was conducted with a focus on evaluating the air pollution distributions within each country. Due to the highly skewed distributions of both population-adjusted dependent variables, namely COVID-19 infections / 100,000 inhabitants, COVID-19 deaths / 100.000 inhabitants, and mortality rates (deaths / infections), we opted for a correlation metric based on ranks instead of raw values. Kendall tau correlation coefficients were employed for all statistical tests.

Because both virus spread and air pollution dynamics present obvious spatially-dependent dynamics, we identified potential clusters of adjacent administrations using Local Moran’s Bivariate statistic^56,57^.

These results are illustrated with thematic maps which better highlight the overlap between air quality and COVID-19 distributions within the eight assessed countries.

## Results

### Correlation between air pollution variables with COVID-19 infections and mortality

Significant positive correlations between air quality variables and COVID-19 infections, fatalities and mortality rate were found in China, the United States, Italy, Iran, France, and the U.K., but not fully in Spain and Germany (Tables 1-3). The strongest correlations were found in Italy, both for infections and deaths, while population size and densities did not explain COVID-19 incidence. In China, population densities showed a similar positive correlation with the virus infections and deaths than air pollution, while in the United States and U.K. population density has a stronger correlation than air pollution variables. In the U.K., air pollution showed a fair degree of correlation with deaths and mortality, but not with the infections. Despite its small sample size (df = 29), Iran shows a significant correlation with NO_2_ distribution and no incidence from population variables. The results for Spain and Germany showed different patterns. The spread of COVID-19 and its related fatalities in Spain could not be explained by differences in air pollution; however, the mortality rate varied with NO_2_ concentration. Moreover, population size and density were negatively correlated with the virus. In a distinct manner, population density explained weakly COVID-19 infections in Germany while the distribution of fine particulate matter was in some cases, weakly negatively correlated. Among the different pollutant analysed, O_3_ and SO_2_ measures from ground stations in China and the United States did not show significant correlations with COVID-19 or were negatively correlated, in contrast with the overall results from the other pollutants.

**Table 1.** Correlation coefficients between COVID-19 infections per 100,000 inhabitants and air quality variables. Significant Kendall correlations (*p*-value < 0.05) are shown in bold; blue and red colour highlights indicate positive and negative correlations, respectively.

|  | China |  |  | U.S.A. |  |  | Italy (provinces) |  |  | Iran |  |  | Spain |  |  | Germany |  |  | U.K. |  |  |
| --- | --- | --- | --- | --- | --- | --- | --- | --- | --- | --- | --- | --- | --- | --- | --- | --- | --- | --- | --- | --- | --- |
|  | df |  | P | df |  | P | df |  | P | df |  | P | df |  | P | df |  | P | df |  | P |
|  | (N-2) | Tau | value | (N-2) | Tau | value | (N-2) | Tau | value | (N-2) | Tau | value | (N-2) | Tau | value | (N-2) | Tau | value | (N-2) | Tau | value |
| pop | 337 | .23 | **<.001** | 3102 | .26 | **<.001** | 105 | .00 | .951 | 29 | -.15 | .250 | 41 | -.27 | **.010** | 399 | .03 | .317 | 362 | .18 | **<.001** |
| pop dens | 337 | .32 | **<.001** | 3102 | .30 | **<.001** | 105 | .12 | .078 | 29 | .14 | .279 | 41 | -.33 | **.002** | 399 | .10 | **.002** | 362 | .21 | **<.001** |
| PM 2.5 sat | 337 | .28 | **<.001** | 3102 | .25 | **<.001** | 105 | .62 | **<.001** | 29 | .24 | .061 | 41 | -.03 | .778 | 399 | -.07 | **.046** | 362 | -.03 | .386 |
| NO<sub>2</sub> sat | 337 | .24 | **<.001** | 3101 | .22 | **<.001** | 105 | .55 | **<.001** | 29 | .40 | **<.001** | 41 | .08 | .470 | 399 | -.03 | .375 | 360 | .06 | .086 |
| PM 2.5 gr | 302 | .15 | **<.001** | 427 | .21 | **<.001** | 88 | .34 | **<.001** |  |  |  |  |  |  |  |  |  |  |  |  |
| PM 10 gr | 302 | .04 | .330 | 201 | .14 | **.004** | 99 | .11 | .096 |  |  |  |  |  |  |  |  |  |  |  |  |
| CO gr | 302 | -.01 | .840 | 156 | .18 | **.001** |  |  |  |  |  |  |  |  |  |  |  |  |  |  |  |
| NO<sub>2</sub> gr | 302 | .12 | **.002** | 246 | .41 | **<.001** |  |  |  |  |  |  |  |  |  |  |  |  |  |  |  |
| O<sub>3</sub> gr | 302 | -.03 | .477 | 749 | .03 | .238 |  |  |  |  |  |  |  |  |  |  |  |  |  |  |  |
| SO<sub>2</sub> gr | 302 | -.01 | .843 | 314 | -.12 | **.002** |  |  |  |  |  |  |  |  |  |  |  |  |  |  |  |

**Table 2.** Correlation coefficients between COVID-19 fatalities per 100,000 inhabitants and air quality variables. Significant Kendall correlations (*p*-value < 0.05) are shown in bold; blue and red colour highlights indicate positive and negative correlations, respectively.

|  | China |  |  | U.S.A. |  |  | Italy (regions) |  |  | France |  |  | Spain |  |  | Germany |  |  | U.K. |  |  |
| --- | --- | --- | --- | --- | --- | --- | --- | --- | --- | --- | --- | --- | --- | --- | --- | --- | --- | --- | --- | --- | --- |
|  | df |  | P | df |  | P | df |  | P | df |  | P | df |  | P | df |  | P | df |  | P |
|  | (N-2) | Tau | value | (N-2) | Tau | value | (N-2) | Tau | value | (N-2) | Tau | value | (N-2) | Tau | value | (N-2) | Tau | value | (N-2) | Tau | value |
| pop | 337 | .17 | **<.001** | 3102 | .36 | **<.001** | 19 | .01 | .976 | 94 | .17 | **.015** | 41 | -.25 | **.019** | 399 | .03 | .409 | 362 | .13 | **<.001** |
| pop dens | 337 | .16 | **<.001** | 3102 | .36 | **<.001** | 19 | .16 | .323 | 94 | .24 | **<.001** | 41 | -.40 | **<.001** | 399 | .05 | .153 | 362 | .29 | **<.001** |
| cases 100k | 337 | .39 | **<.001** | 3102 | .55 | **<.001** | 19 | .83 | **<.001** |  |  |  | 41 | .81 | **<.001** | 399 | .65 | **<.001** | 362 | .46 | **<.001** |
| PM 2.5 sat | 337 | .18 | **<.001** | 3102 | .24 | **<.001** | 19 | .60 | **<.001** | 94 | .56 | **<.001** | 41 | -.09 | .385 | 399 | -.04 | .241 | 362 | .16 | **<.001** |
| NO<sub>2</sub> sat | 337 | .16 | **<.001** | 3101 | .26 | **<.001** | 19 | .51 | **<.001** | 94 | .57 | **<.001** | 41 | .08 | .470 | 399 | -.05 | .118 | 360 | .23 | **<.001** |
| PM 2.5 gr | 302 | .18 | **<.001** | 427 | .24 | **<.001** | 17 | .22 | .183 |  |  |  |  |  |  |  |  |  |  |  |  |
| PM 10 gr | 302 | .12 | **.006** | 201 | .18 | **<.001** | 19 | .00 | 1.00 |  |  |  |  |  |  |  |  |  |  |  |  |
| CO gr | 302 | .11 | **.012** | 156 | .20 | **<.001** |  |  |  |  |  |  |  |  |  |  |  |  |  |  |  |
| NO<sub>2</sub> gr | 302 | .12 | **.005** | 246 | .42 | **<.001** |  |  |  |  |  |  |  |  |  |  |  |  |  |  |  |
| O<sub>3</sub> gr | 302 | -.02 | .585 | 749 | .03 | .173 |  |  |  |  |  |  |  |  |  |  |  |  |  |  |  |
| SO<sub>2</sub> gr | 302 | .04 | .409 | 314 | -.08 | **.028** |  |  |  |  |  |  |  |  |  |  |  |  |  |  |  |

**Table 3.**
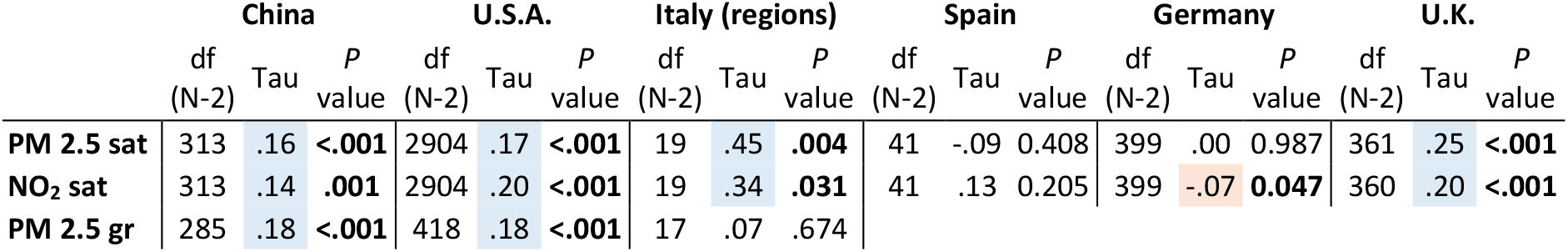
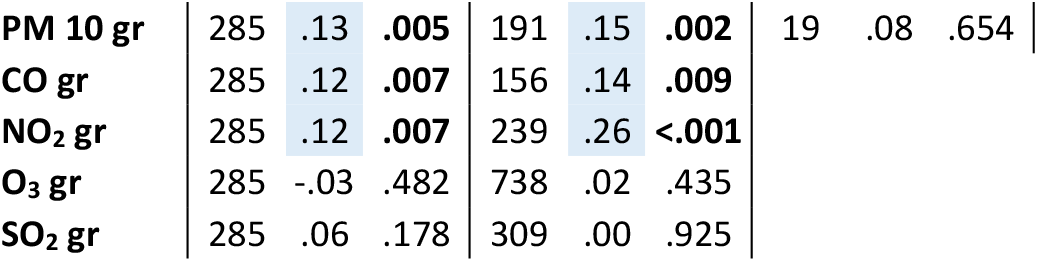
Correlation coefficients between COVID-19 mortality rates and air quality variables. Significant Kendall correlations (*p*-value < 0.05) are shown in bold; blue and red colour highlights indicate positive and negative correlations, respectively.

|  | China |  |  | U.S.A. |  |  | Italy (regions) |  |  | Spain |  |  | Germany |  |  | U.K. |  |  |
| --- | --- | --- | --- | --- | --- | --- | --- | --- | --- | --- | --- | --- | --- | --- | --- | --- | --- | --- |
|  | df |  | P | df |  | P | df |  | P | df |  | P | df |  | P | df |  | P |
|  | (N-2) | Tau | value | (N-2) | Tau | value | (N-2) | Tau | value | (N-2) | Tau | value | (N-2) | Tau | value | (N-2) | Tau | value |
| PM 2.5 sat | 313 | .16 | **<.001** | 2904 | .17 | **<.001** | 19 | .45 | **.004** | 41 | -.09 | .408 | 399 | .00 | .987 | 361 | .25 | **<.001** |
| NO<sub>2</sub> sat | 313 | .14 | **.001** | 2904 | .20 | **<.001** | 19 | .34 | **.031** | 41 | .13 | .205 | 399 | -.07 | **.047** | 360 | .20 | **<.001** |
| PM 2.5 gr | 285 | .18 | **<.001** | 418 | .18 | **<.001** | 17 | .07 | .674 |  |  |  |  |  |  |  |  |  |

|  |  |  |  |  |  |  |  |  |  |
| --- | --- | --- | --- | --- | --- | --- | --- | --- | --- |
| PM 10 gr | 285 | **.13** | **.005** | 191 | **.15** | **.002** | 19 | .08 | .654 |
| CO gr | 285 | **.12** | **.007** | 156 | **.14** | **.009** |  |  |  |
| NO<sub>2</sub> gr | 285 | **.12** | **.007** | 239 | **.26** | **<.001** |  |  |  |
| O<sub>3</sub> gr | 285 | -.03 | .482 | 738 | .02 | .435 |  |  |  |
| SO<sub>2</sub> gr | 285 | .06 | .178 | 309 | .00 | .925 |  |  |  |

### COVID-19 distribution, clusters and air quality maps

Figure 1 reports comparison maps of COVID-19 distributions with the satellite-based PM 2.5 concentrations for the eight analysed countries. These graphical representations also allow for a rapid assessment of the air pollution pattern in each country (basic descriptive statistics for each pollutant can be found in S.I., Table S2). While the PM 2.5 maps are continuous surfaces drawn following the same classification scheme across countries, the COVID-19 infections and fatalities maps required ad-hoc classification adaptations due to different population profiles and infection dynamics. In China, due to the very large population and an apparently effective policy for the containment of the virus, the number of cases per 100,000 residents were relatively low and highly concentrated in the epicentre of the outbreak (Wuhan and the Hubei province). A visual correlation between the two maps can be perceived, especially between the eastern and western parts of the country, and it’s clearly highlighted in the cluster map (Figure 2). The highly developed and polluted areas in the east of China represent outlier clusters due to the low COVID-19 cases compared to the Hubei province. In the United States, the virus noticeably appears to spread over several areas. PM 2.5 differences are not large, but their distribution looks adequately coincident with the fatalities. Similar to China, a longitudinal pattern is visible with low-fatalities / low-pollution clusters (LL) concentrated in the mid-western part of the country while high-fatality and high-pollution clusters (HH) are found in the east, along the Mississippi river and the states surrounding New York. However, a high number of outliers of both types (HL and LH) exist. The high correlation results found for Italy are clearly visible. The polluted areas of the Po valley are those heavily affected by COVID-19 infections. The clusters are clear and the number of outliers is very limited. While in Iran and France the correlations are only lightly perceivable, the clusters maps show a north-south regionalization pattern similar to Italy. The maps of Spain confirm the absence or weak correlation shown in Tables 1-3, apparently going against our general hypotheses. Nevertheless, PM 2.5 levels in Spain are minimal, as well as their variation, as indicated by the low range and inter-quartile range (Table S2 in S.I.). The U.K. map of PM 2.5 shows well the higher concentrations around urban areas and the overall south-eastern area where COVID-19 mortality is higher too. However, also a few counties/NHS in the north of Scotland are particularly affected by the virus infections, becoming outliers in the clusters map. Finally, COVID-19 mortality in Germany is low and no apparent distribution pattern can be detected, being quite well spread. Similarly, PM 2.5 concentrations are fairly high all over the country, with peaks in the eastern districts, where a few HH clusters and LH outliers are found. The high number of non-significant clusters and both types of outliers confirm this tendency to a homogenous distribution of COVID-19 and air pollution.

**Figure 1.**
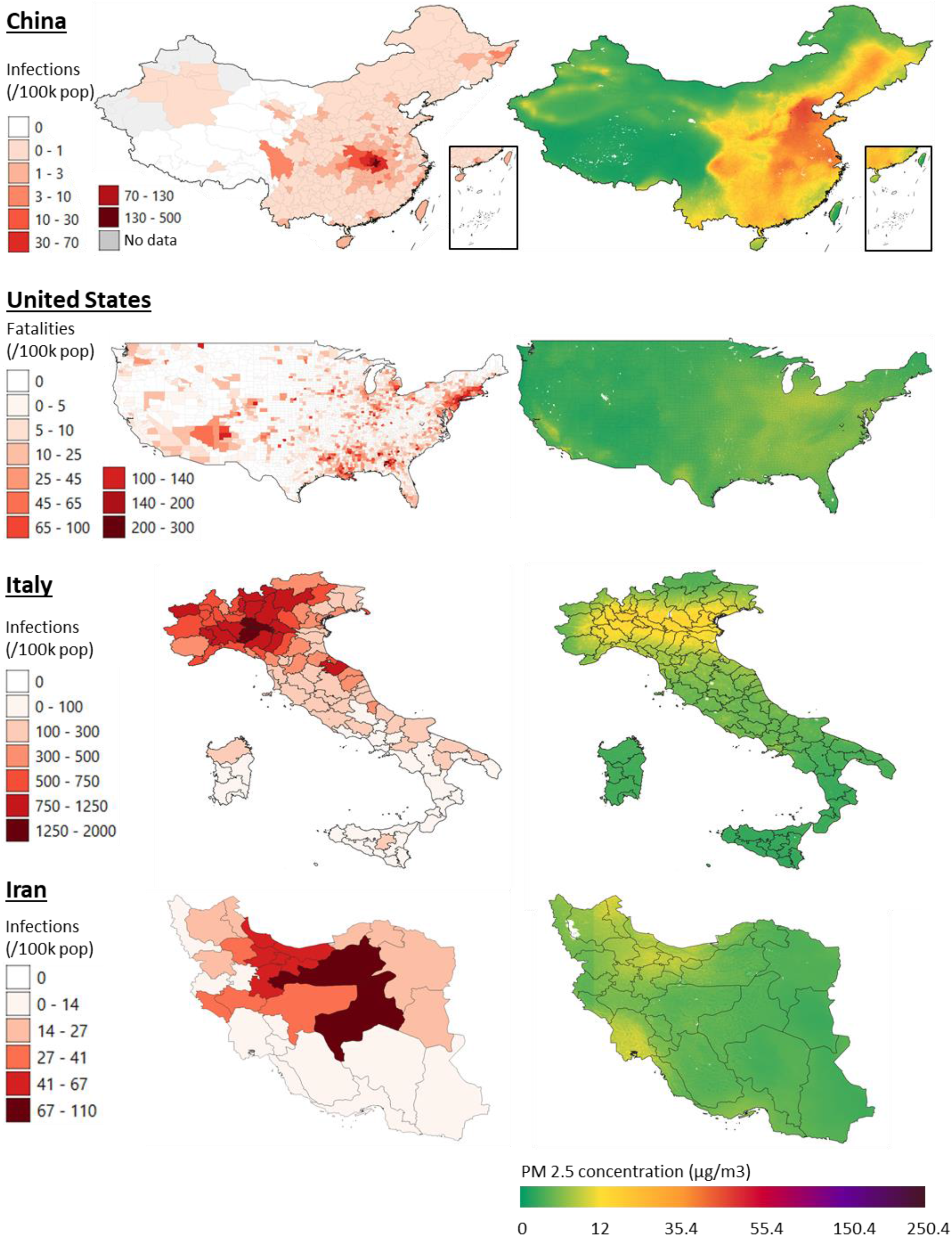
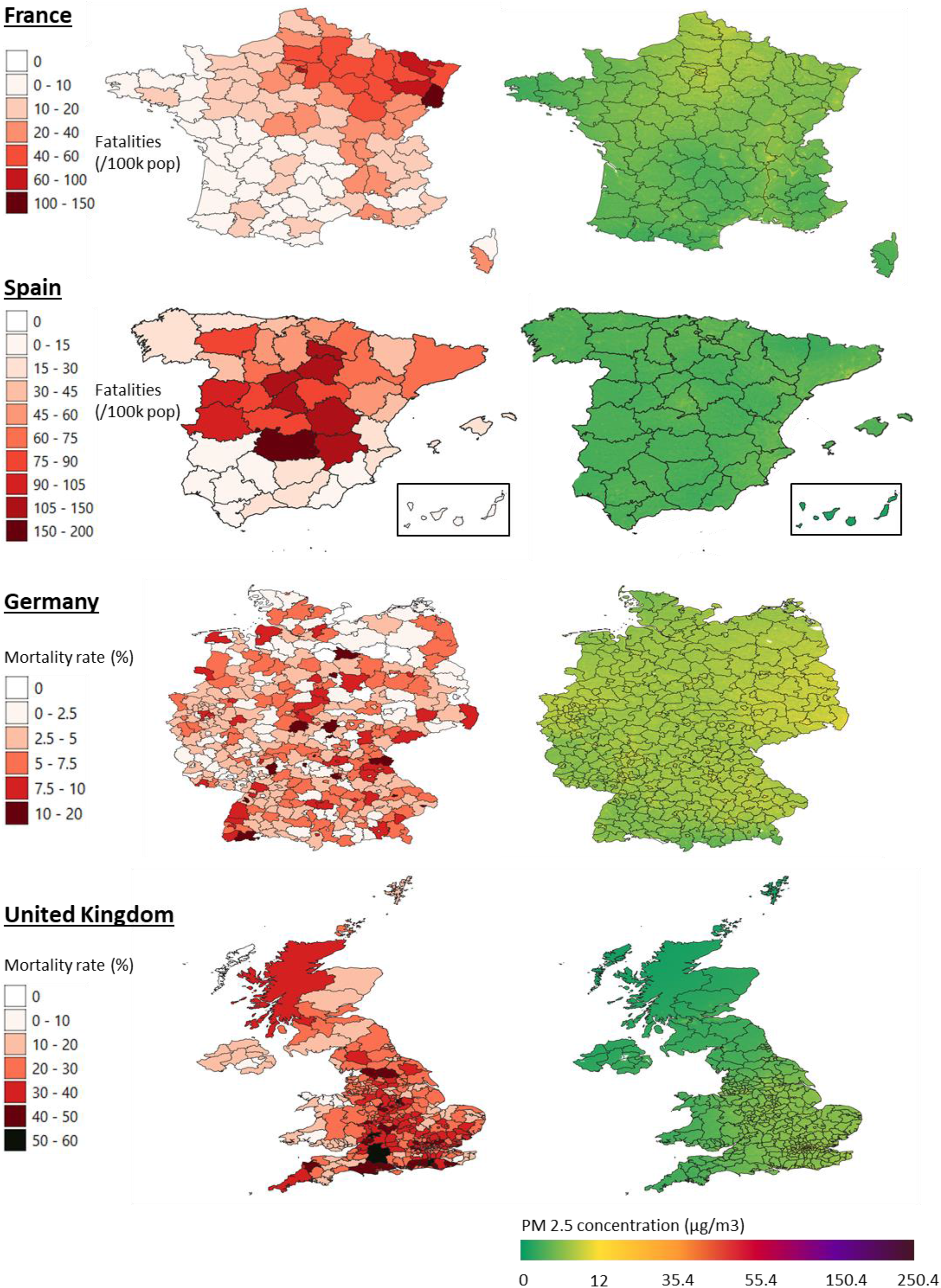
Maps comparisons of satellite-derived PM 2.5 distributions and COVID-19 infections, fatalities (per 100,000 inhabitants), or mortality rates (deaths / infections) in eight countries (see S.I. for information about dates). Maps of different countries should not be compared due to different classification schemes and spatial scales. Administrative units’ boundaries were seldom adapted to the COVID-19 data available (e.g. merged districts of Galicia, Catalunia and Pais Vasco in Spain).

**Figure 2.**
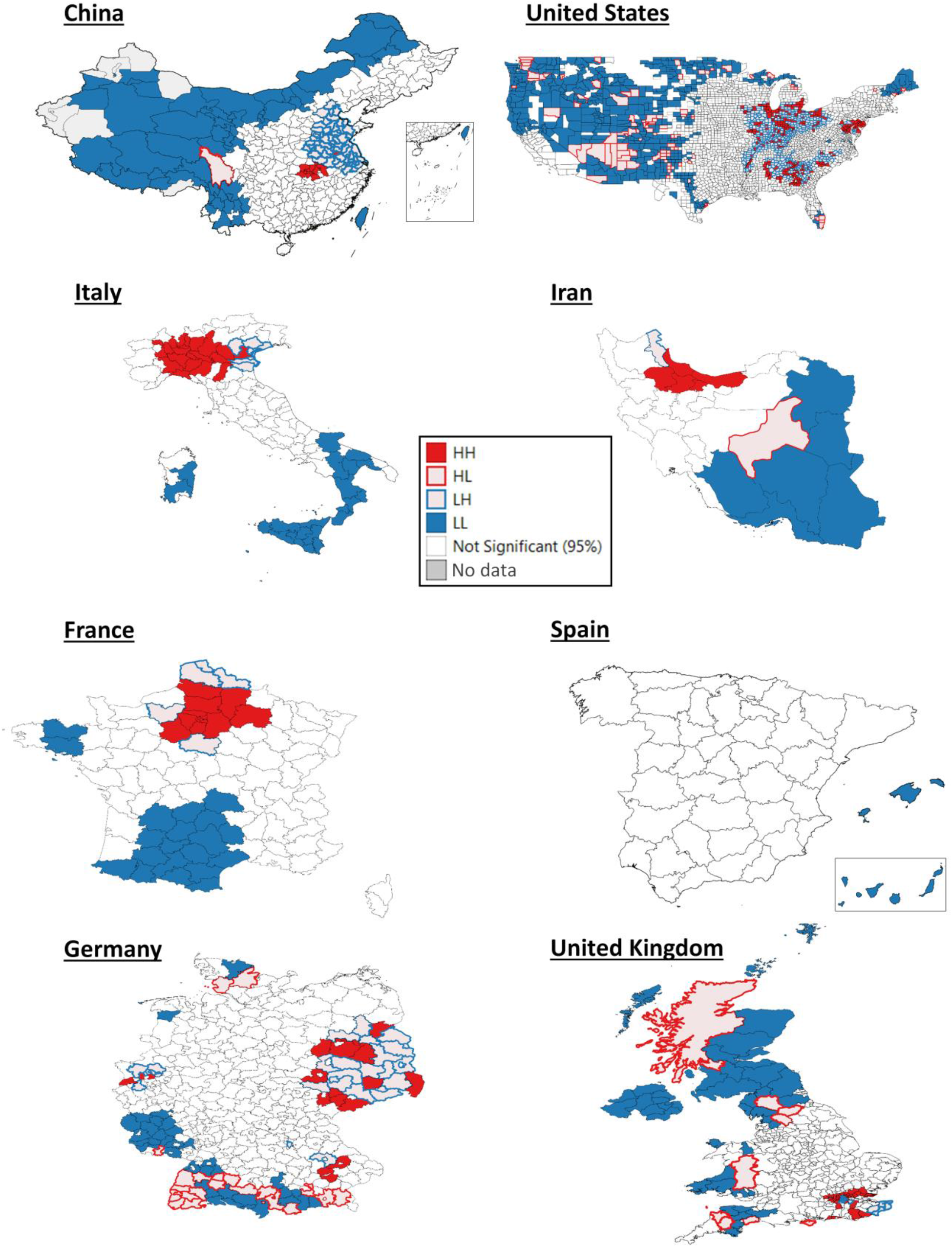
Maps of clustered, adjacent administrations resulting from Local Moran Bivariate analysis. Four types of significant spatial relationship exist. HH in filled red: High COVID-19 (fatalities, deaths, or mortality rates) and High PM2.5; HL in empty red: High COVID-19 and Low PM2.5; LH in empty blue: Low COVID-19 and High PM2.5; LL in filled blue: Low COVID-19 and Low PM2.5. While HH and LL filled coloured clusters support the COVID-19/air pollution correlation hypothesis, HL and LH empty colours represent outliers. Note that the COVID-19 variables used in each country are those of Figure 1.

## Discussion

This study is the first to investigate long-term air pollution exposure measured by satellite and ground sensors for eight countries as a potential and highly likely risk factor for the incidence and mortality rates of COVID-19. It provides evidence that SARS-CoV-2 infections are most often found in highly polluted and densely populated areas. In some cases (Italy and Iran), air pollution independent from population density explained the distribution pattern of the virus. In addition, in these areas affected by a mixture of air pollutants, the virus kills more frequently than elsewhere. In the questionable case that the figures provided by these eight states in relation to the number of infections and deaths are inaccurate^58^, our analyses and conclusions would not need to be reframed. In fact, if that was the case, this error would most likely be concentrated in just one or very few administrations, or it would be evenly spread across administrations, not affecting the general significance of the correlations.

In Chinese cities^59^ and, more in detail, in the Hubei province, **brand new** time analyses give preliminary evidence of a correlation between high levels of NO_2_ and 12-day delayed virus outbreaks^60^ and other PM covariates^61,62^. With this paper, we therefore add the long-term exposure effects for China. As shown in the maps, China bears extremely high rates of air pollution, concentrated in the east. However, COVID-19 cases occurred mainly in the constrained area of Hubei. All evidence suggests that the enforced lockdown was the major factor controlling the virus spread. It is peculiar that the onset of the pandemic still appeared in one of the most polluted areas of the globe…

In the **United States**, an increase of a mere 1 μg/m^3^ in PM 2.5 was recently found responsible for an 8% higher mortality rate by COVID-19, a rate relatively higher than other 11 demographic co-variables tested^63^. With our paper, we add PM 10, NO_2_, and CO measures from ground stations. Our analysis for the United States was run over more than 3000 administrative units, many of which were still relatively unaffected by the virus (as of 21 May 2020), despite being the country featuring the highest COVID-19 numbers.

We found that in **Italy**, the correspondence between poor air quality and SARS-CoV-2 appearance as well as its induced mortality was the starkest. The area with the largest number of infections and deaths in Italy is the Po Valley, which is also the foremost place of polluted air in Europe^64^. This result was first hypothesised^65^ and later confirmed by another study^66^ and a remarkable further one^67^ which included five demographic co-variables.. The fact the population density does not play a role in the incidence of COVID-19 in Italy and Iran is a unique finding in our investigation, which strongly supports the common hypotheses of all these other, similar studies, and questions the common criticism, stating that air polluted areas are also those more densely populated (see table S3). Other factors must be attributed for such a severe incidence in Italy, including its very large aging population. Not to forget that the aged population is also the one which got exposed to air pollutants for the longest time. Such pollutants cause, in turn, other comorbidities and COVID-19 vulnerabilities such as cardiocirculatory diseases.

Unfortunately, information on COVID-19 infections were available for **Iran** at the first administration level only till 22 March 2020. **France** provides up-to-date information about fatalities only. Despite these limitations, Iran and France show highly clustered COVID-19/pollution distributions, in the same manner as the Italian case, resulting in significant positive correlation that confirm our hypotheses.

The absence of correlation found in **Spain** should be attributed to high levels of air quality, within the green Air Quality Index standard range, and ensuing minimal differences among its provinces. Moreover, the regions most affected by the virus seem to be those less densely populated, a peculiarity still not provided by the literature and requiring future investigation. In **Germany**, also, a clear correlation could not be detected because, inversely, pollution is widely spread across its districts. For these two countries, we therefore back up another report^68^ which finds high levels of NO_2_ associated with COVID-19 mortality in Spain and Germany. To note, though, that it is always necessary to control for population densities in these types of analysis^69^.

Finally, in the **U**.**K**., where containment measures were implemented late, fatalities and mortality rates, but not cases alone, were correlated with air pollution, suggesting that when affected by the disease, a weakened respiratory system due to prolonged stress by air pollution increases the risk of mortality.

Despite the highly significant correlations of these findings that we collected over three time periods in March^54^, April^70^ and, as reported here, end of May, their interpretation has to be necessary **cautious**. The virus spread in most countries is still ongoing^71^ and is being contained^44^. Causation should not be inferred by correlational data alone. There are also confounding factors such as how the virus infection was determined in patients by different countries. However, the larger the geographical areas are affected by the pandemic, the lower these elements play a role. Finally, it should be noted that by accounting for yearly averaged air quality indexes, we accounted for the long-term exposure to these pollutants, therefore keeping on the conservative side. These correlations would become even stronger, in fact, when limiting the analysis to the more polluted winter months, given how they invariably bear lower air quality.

We run these analyses considering eight countries in their second-order administrations’ level. If controlling for several **other predictors** like demographic variables is something advisable to perform at a single-country level, so to cross-check for the interdependence with other predictors, including them at an international scale poses an apparent technical limitation^72^. National or federal health systems have different capacities and provide care in their distinct ways. This in turn influences case detections, intensive care capacity and fatality rates. Cofactors such as earliest location of the pathogen, population mobility and patient socioeconomic status or ethnicity may not be accounted reliably between such diverse countries spanning from Asia to the western world, because they are interdependent and only in part nested within countries or administrations, even when included as random factors as in a comprehensive generalized mixed model. Yet the epidemics, which has turned into a pandemic, might have catered for this limitation; the larger its extent, the more prominent a common factor like air pollution has become, while other secondary predictors will level out across places.

Three **additional countries** were reported to follow this same trend between PM 2.5 and the virus: the Netherlands^73^, controlling for some other medical risk factors; Japan^74^, finding a positive correlation in the elderly; and also India, which holds a similar trend (Sakthi J. Sundar R., in prep.). To note, finally, that CO_2_ and SO emissions keep being correlated with this disease throughout 126 countries when analysed with the “Our World in Data” database^75^. Ambient, outdoor air pollution, causing an estimated 4.2 million deaths yearly worldwide^76^, is indeed a worthwhile risk cofactor to be hypothesized in connection with a new respiratory disease, without necessarily having to analyse some of the other cofactors.

Since there is now some first evidence that the cross of the virus from animals to humans may have happened earlier than the end of 2019^77^ and further south than in the Chinese city of Wuhan^78^, we can speculate that air pollution could have allowed the new epidemics to become recognised due to an influx of patients with weak respiratory systems showing higher morbidity and mortality than influenza. The same seems to have happened in Europe, since the virus moved from central Europe to the most polluted region of the continent, in northern Italy^44,79^.

**Further research** in the field of genetics will ascertain whether virulence has evolved in the air polluted places of those countries where a gradient of air pollution is present. The initial location of the pathogen, travelling and super-spread events are deemed to be foremost factors governing the epidemics (further discussed in S.I.). Later on, other factors such as pollution may become major predictors for harsh infections to manifest. In relation to short-term exposure to peaks of pollutions, the capacity of air pollutants to act as viral vectors should be investigated further. Particulate matter in fact does act as a medium for the aerial transport of SARS-CoV-2^39,80^. Aggregates of particulate matter with this virus have been collected in the worst affected northern Italian city of Bergamo^81^. If the viral load carried by the aggregates is enough to cause morbidity, pollution would directly act as a vector, broadening the harm done by the human to human contagions.

To conclude, these findings are sufficiently significant to encourage future research, to always consider air pollution as a contributing risk factor for COVID-19. These results inform epidemiologists and **policy** makers on how to prevent future, more frequent and lethal viral outbreaks by curbing air pollution and, ultimately, meeting climate goals^82^. Can the fossil fuel economy really carry on unabated once we resume the lockdowns? Institutions need to endorse these interventions and speed up reforms more seriously^83-85^, together with endorsing collateral and more comprehensive measures^86^ playing a role in epidemics and zoonoses^87,88^, such as impeding biodiversity loss^89-92^, decreasing intensive livestock farming, and alleviating poverty^93,94^. This new coronavirus shall be an opportunity given to the governments to revive sustainable development goals.

## Acknowledgements

Lei Shi and Xiao Wen commented on the study. Livia Ottisova improved and revised the manuscript. Chun Chen and Michele Mignini commented on statistics. Rafael Moreno Ripoll and Mehrdad Samavati helped to obtain the data for Spain and Iran. RP did this work while in isolation in the Italian Po valley, due to the ongoing pandemic.

## Data Availability

The compiled datasets are available at https://github.com/DavideFornacca/COVID19/tree/master/8_countries.

## Author Contributions

Concept and design: RP.

Data acquisition and statistical analysis: DF.

Interpretation: DF and RP.

Drafting of the manuscript: RP.

Critical revision of the manuscript: RP and DF.

## Supporting information

### Supplement to Introduction

Several **risk factors** have been implicated with the fast spread of SARS-CoV-2, including super spread events^1,2^. Its further spread to different countries has been attributed to air travellers^3-11^. A number of personal risk factors have further been implicated with higher morbidity and mortality rates of Covid-19, including male gender and smoking status. In particular, smoking has been associated with a higher morbidity and mortality of COVID-19 in men than in women^12^.

The temperate-climate **latitudes** have been identified as the probable areas to be mostly affected by COVID-19^13^ due to a limited exposure to UV light in winter. The sole temperature^14,15^ or humidity^16^ appear to play less of a role^17^. Indeed, other human coronaviruses (HCoV-229E, HCoV-HKU1, HCoV-NL63, and HCoV-OC43) appear between December and April, and are undetectable in summer months in temperate regions, leading to winter seasonality behaviour.

The very **first appearance** of this virus cannot be directly correlated with one of these predictors, since, like the other SARS coronaviruses, SARS-CoV-2 is alleged to have transferred host from the originating bats to humans^18^. However, it still appeared in a Chinese area affected by some of the highest air pollution in the world, and it showed a relatively high virulence there.

### Supplement to Methods

We selected the eight countries according to the following elements. China (including Taiwan, Hong Kong and Macau) was chosen because of its large size and now advanced stage of its epidemic, well into the recovery phase. The second choice was Italy, at the time of writing among the most heavily affected countries of the world, and just passed the peak of contagion. The area with the largest number of infections and deaths in Italy is the Po Valley, which is also the foremost place of polluted air in Europe^19^. The third country investigated was the conterminous United States, which currently has the highest number of cases worldwide, yet is still behind in the pandemic curve due to its later arrival as compared to Asia and Europe. Among the countries where the virus spread earlier, we included Iran which heavily suffers from severe air pollution due to the ubiquitous use of gas methane, refineries and heavy traffic. France and Spain were selected because of the high COVID-19 figures, but minor air pollution issues than Italy. Lastly, Germany and the United Kingdom represented suitable candidates to feed into the analysis, because of the relatively reduced lockdown measures adopted by those states^20^.

For Iran, the state made public the data of infections at a first level resolution only, until 22 March 2020. The Chinese dataset includes the 17 April update with a 50% increase in deaths in Wuhan city^21^. Fatalities in Italy were available only at the regional level, therefore two different datasets were compiled. The autonomous communities of Catalonia, Galicia, and Pais Vasco in Spain provided figures at the first administration level only^22^, so we considered them at the same level as provinces. For France, COVID-19 deaths only were available at the department level. Finally, in the U.K., the data for Scotland was organized following the National Health Service (NHS) subdivisions rather than the second order administration scale.

Global Annual PM 2.5, Grids from MODIS, MISR and SeaWiFS Aerosol Optical Depth (AOD) with GWR, v1 (1998 – 2016), were obtained from NASA’s Socioeconomic Data and Applications Center^23,24^. From the same repository, we retrieved the Global 3-Year Running Mean, Ground-Level NO2 Grids from GOME, SCIAMACHY and GOME-2, v1 (1996 – 2012)^23,24^.

For both products, the annual grids were first reduced to an average multi-year image and afterwards, the mean of all grid cells covering every administrative unit was calculated.

The latest available annual means of measured PM 2.5 and PM 10 values were retrieved from the World Health Organization Global Ambient Air Quality Database of 2018^25^ and used for the Italian case study because of extensive coverage. For China, we used aggregated monthly air quality data for the years 2014 to 2016, made available by the Center for Geographic Analysis Dataverse of the University of Harvard^26^. For the conterminous U.S.A., summary data on several pollutants for the year 2019 was retrieved from the United States Environmental Protection Agency^27^. To every administrative unit, we assigned the air quality value from its related station. If more than one point fell within a given unit, the mean was calculated. No ground measures for the other countries were included in our study.

**Table S1.**
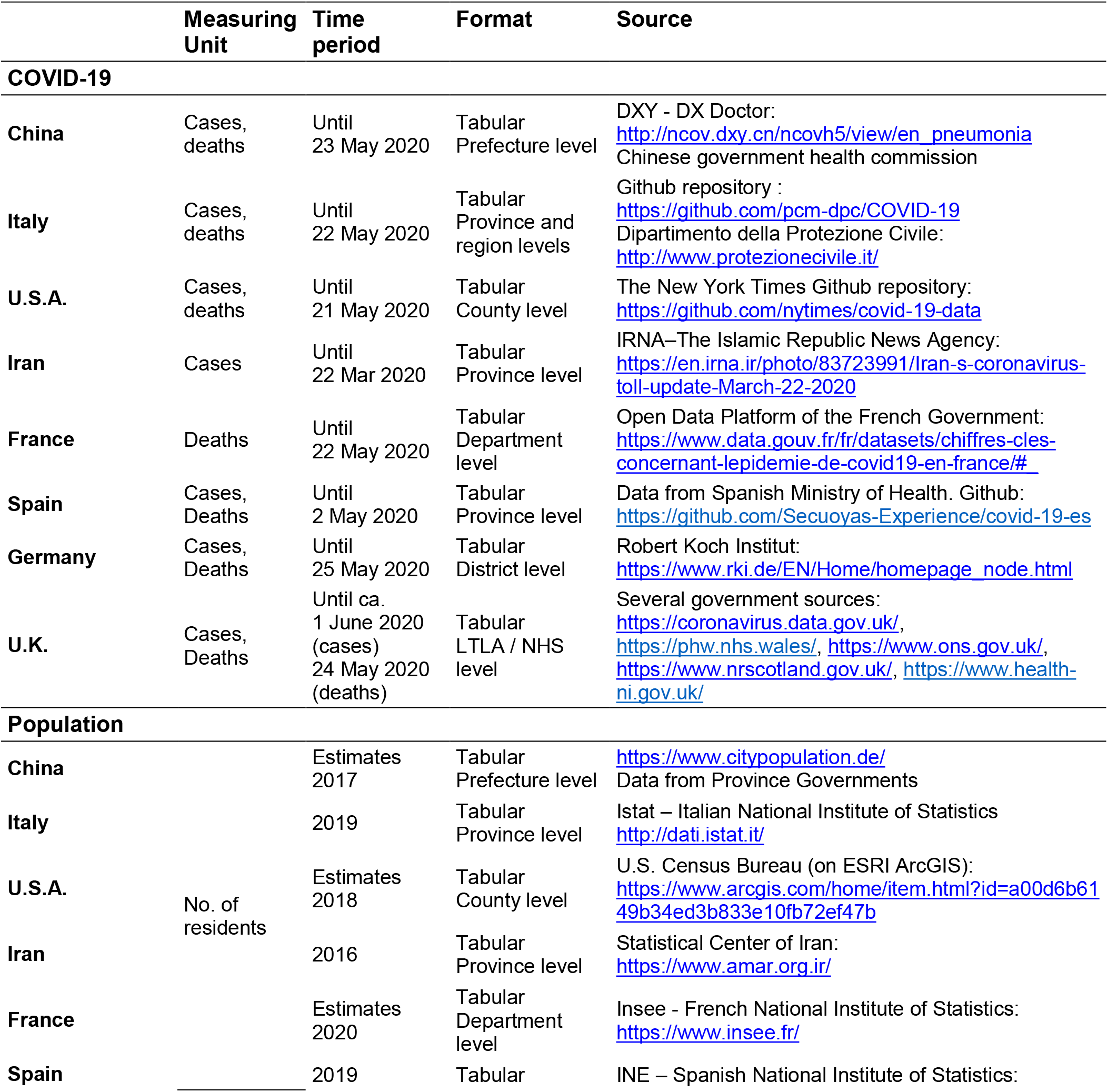
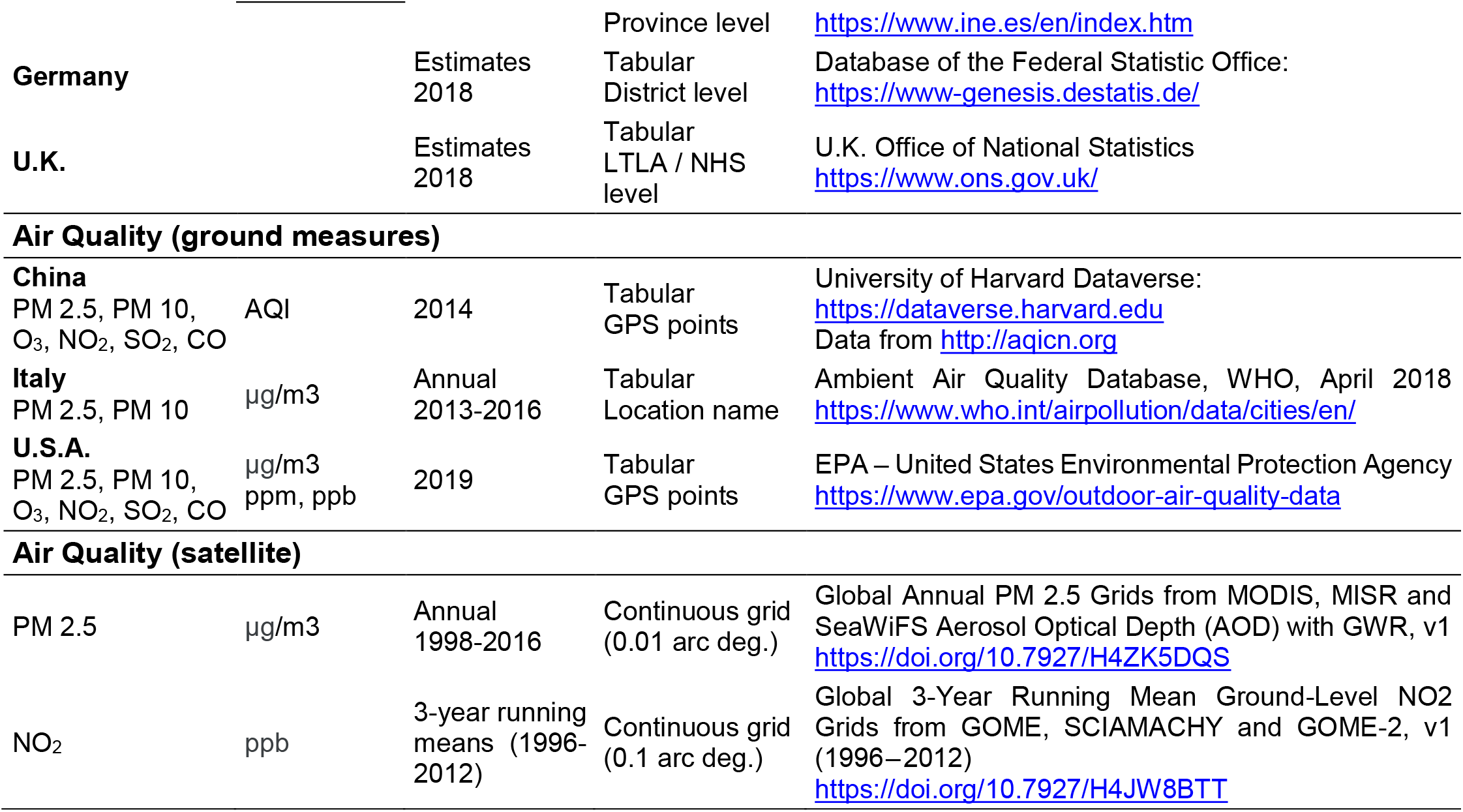
Detailed information on the datasets used for the viral and pollution analyses.

### Supplement to Results

**Table S2.** Descriptive statistics for the air pollution variables in the eight analysed countries.

|  | variable | unit | count | mean | std | min | 25% | 50% | 75% | max | range | iqr |
| --- | --- | --- | --- | --- | --- | --- | --- | --- | --- | --- | --- | --- |
| China | PM25_sat | ug/m3 | 347 | 30.31 | 15.80 | 2.08 | 19.11 | 29.09 | 40.01 | 70.98 | 68.90 | 20.89 |
|  | NO2_sat | ppb | 347 | 1.96 | 1.96 | 0.06 | 0.54 | 1.26 | 3.06 | 13.75 | 13.69 | 2.52 |
|  | PM25_gr | AQI | 308 | 110.78 | 27.79 | 38.20 | 92.63 | 111.55 | 128.51 | 186.96 | 148.76 | 35.88 |
|  | PM10_gr | AQI | 308 | 63.61 | 22.98 | 19.96 | 46.26 | 60.90 | 75.67 | 170.27 | 150.31 | 29.40 |
|  | CO_gr | AQI | 308 | 9.71 | 3.97 | 2.39 | 7.11 | 8.74 | 11.85 | 27.01 | 24.63 | 4.74 |
|  | NO2_gr | AQI | 308 | 13.72 | 5.40 | 3.17 | 9.63 | 13.40 | 17.41 | 28.57 | 25.40 | 7.78 |
|  | O3_gr | AQI | 308 | 25.83 | 5.87 | 14.22 | 21.70 | 25.17 | 28.98 | 50.54 | 36.31 | 7.28 |
|  | SO2_gr | AQI | 308 | 13.24 | 7.96 | 1.14 | 7.88 | 10.97 | 16.46 | 40.58 | 39.44 | 8.58 |
| U.S.A. | PM25_sat | ug/m3 | 3104 | 9.37 | 2.68 | 2.32 | 7.26 | 9.68 | 11.54 | 15.57 | 13.24 | 4.28 |
|  | NO2_sat | ppb | 3103 | 1.59 | 1.25 | 0.17 | 0.73 | 1.20 | 2.13 | 14.97 | 14.80 | 1.40 |
|  | PM25_gr | ug/m3 | 429 | 7.22 | 2.10 | 0.00 | 5.88 | 7.37 | 8.66 | 15.73 | 15.73 | 2.78 |
|  | PM10_gr | ug/m3 | 203 | 16.09 | 6.36 | 4.60 | 12.41 | 15.43 | 18.62 | 40.64 | 36.04 | 6.21 |
|  | CO_gr | ppm | 158 | 0.25 | 0.10 | 0.04 | 0.19 | 0.25 | 0.30 | 0.82 | 0.78 | 0.12 |
|  | NO2_gr | ppb | 248 | 14.63 | 7.93 | 1.08 | 7.88 | 14.50 | 20.65 | 36.73 | 35.65 | 12.76 |
|  | O3_gr | ppm | 751 | 0.05 | 0.00 | 0.03 | 0.04 | 0.05 | 0.05 | 0.06 | 0.03 | 0.00 |
|  | SO2_gr | ppb | 316 | 2.61 | 5.56 | -0.38 | 0.57 | 1.30 | 2.51 | 75.47 | 75.85 | 1.94 |
| Italy<br>(provinces) | PM25_sat | ug/m3 | 107 | 12.82 | 6.08 | 4.50 | 7.68 | 11.62 | 18.10 | 25.37 | 20.86 | 10.42 |
|  | NO2_sat | ppb | 107 | 2.67 | 2.42 | 0.39 | 0.99 | 1.61 | 3.95 | 11.56 | 11.16 | 2.96 |
|  | PM25_gr | ug/m3 | 90 | 16.37 | 4.89 | 6.00 | 13.00 | 15.37 | 19.46 | 29.00 | 23.00 | 6.46 |
|  | PM10_gr | ug/m3 | 101 | 23.67 | 5.61 | 13.67 | 19.75 | 22.50 | 26.75 | 41.00 | 27.33 | 7.00 |
| Italy<br>(regions) | PM25_sat | ug/m3 | 21 | 11.29 | 4.73 | 4.90 | 7.73 | 10.48 | 13.84 | 20.59 | 15.69 | 6.11 |
|  | NO2_sat | ppb | 21 | 1.93 | 1.68 | 0.48 | 0.83 | 1.35 | 2.25 | 7.08 | 6.59 | 1.42 |
|  | PM25_gr | ug/m3 | 19 | 15.19 | 3.50 | 9.50 | 13.00 | 14.67 | 16.05 | 22.96 | 13.46 | 3.05 |
|  | PM10_gr | ug/m3 | 21 | 22.15 | 4.40 | 15.60 | 20.40 | 21.29 | 22.92 | 34.67 | 19.07 | 2.52 |
| Iran | PM25_sat | ug/m3 | 31 | 10.97 | 3.07 | 5.89 | 8.74 | 10.47 | 13.74 | 16.43 | 10.54 | 5.00 |
|  | NO2_sat | ppb | 31 | 0.47 | 0.47 | 0.10 | 0.22 | 0.31 | 0.42 | 2.24 | 2.14 | 0.21 |
| France | PM25_sat | ug/m3 | 96 | 10.23 | 2.42 | 6.22 | 8.13 | 9.90 | 11.79 | 16.49 | 10.27 | 3.66 |
|  | NO2_sat | ppb | 96 | 2.37 | 1.75 | 0.55 | 1.20 | 1.80 | 3.02 | 8.79 | 8.24 | 1.82 |
| Spain | PM25_sat | ug/m3 | 43 | 7.05 | 1.13 | 2.36 | 6.49 | 6.93 | 7.54 | 10.10 | 7.74 | 1.05 |
|  | NO2_sat | ppb | 43 | 1.02 | 0.45 | 0.10 | 0.69 | 0.91 | 1.27 | 2.69 | 2.58 | 0.58 |
| Germany | PM25_sat | ug/m3 | 401 | 13.95 | 1.53 | 8.91 | 13.10 | 13.86 | 14.93 | 18.47 | 9.56 | 1.83 |
|  | NO2_sat | ppb | 401 | 4.77 | 2.04 | 1.48 | 3.48 | 4.19 | 5.24 | 11.86 | 10.38 | 1.76 |
| U.K. | PM25_sat | ug/m3 | 364 | 10.51 | 2.64 | 2.16 | 9.06 | 11.29 | 12.07 | 15.37 | 13.21 | 3.01 |
|  | NO2_sat | ppb | 362 | 5.47 | 2.24 | 0.48 | 4.12 | 5.26 | 6.62 | 10.23 | 9.76 | 2.50 |

**Table S3.**
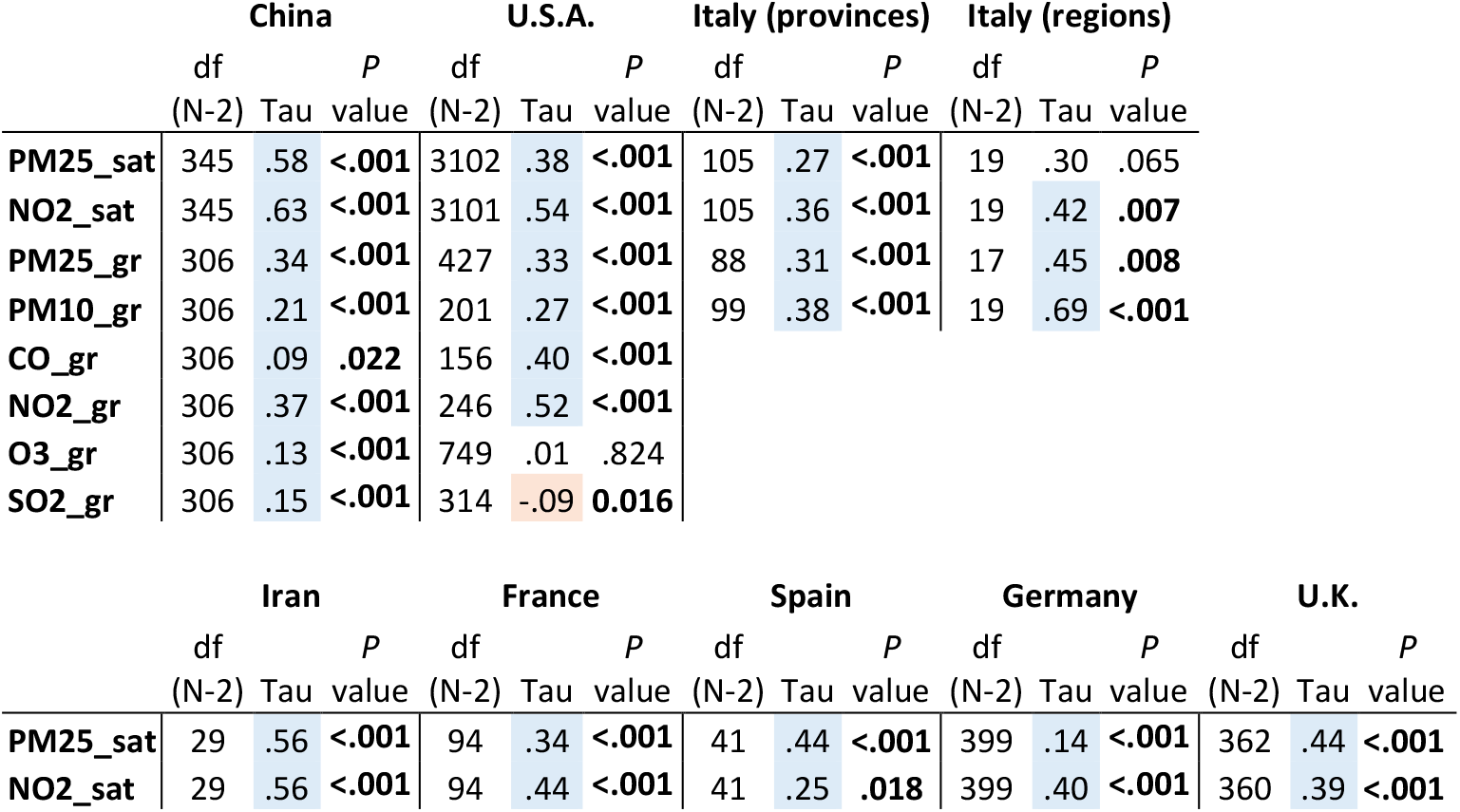
Correlation coefficients between population density and air pollution variables in the eight analysed countries. Significant correlations (*p-*value < 0.05) are shown in bold; blue and red colour highlights indicate positive and negative correlations, respectively.

### Supplement to Discussion

The possibly large proportion of **asymptomatic cases** has been implied as an important factor in the fast spread of the virus and will necessarily lead to a biased mortality rate. Different government policies with regards to testing have led to vastly different estimates across countries^28,10^, and a COVID-19 overall mortality rate has not been established yet. Asymptomatic cases could be as high as about 50% of total cases, as estimated by simulations^29^.

**Satellite NO**_**2**_ **and PM 2**.**5** on their own are suitable representatives for general air quality^30^, since they are more consistent than ground station data, providing finer detail, consistency, accuracy, with virtually no errors of counterfeits. Despite limited by cloud cover, they are to a certain extent less prone to biases from other conditions^14^ of wind and greenhouse effect of temperature inversion, in turn also related to air pollution. In spite of this, we found some consistency between ground and satellite data for those countries for which we could obtain fine-grained ground measures. The satellite databases employed in this study represent annual means, which we further averaged over the full time-series. This only partially represents the real emission of pollutants during the year and do not make evident seasonal variations and other fluctuations. However, our aim was to highlight differences in air quality within a country’s region and show the correlation with the virus. Therefore, threshold values of air pollution cannot be inferred from this study.

States have responded by trying to mitigate the spread of the virus through imposing widespread lockdowns. This has led to a **decrease in air pollution**^31-35^, which in the top faring country in this regard, China, it prevented the equivalent savings of CO_2_ emitted in one year by France^36^, and it likely prevented the deaths of 4,000 children under 5 and 73,000 adults over 70^37^. However, the winter months and low temperature caused people to keep at least domestic heating systems on, maintaining a certain amount of emissions. In Europe^38^ and in China^39^ a consistent reduction in air pollution was recorded by satellites due to reduced anthropogenic activities during the lockdowns, although it occurred gradually^40,41^, also due to weather conditions unfavourable to air quality. Forest fires also decreased by 4.54%^42^.

The quarantines certainly decreased the role that **commuting** has in the virus spread. Nonetheless, reduced anthropogenic activities and reduced mobility lose correlational significance over time, after the first stages of the infection^43,44^. Instead, the correlation we found with low air quality remains significant throughout the different epidemic stages.

